# MR-based normative volumetric analyses reveal genotype-dependent brain anomalies in *FGFR*-related craniosynostoses

**DOI:** 10.64898/2026.11.14.26367821

**Authors:** Ombline Delassus, Jérémy Sadoine, Giovanna Paternoster, Lucas Chollet, Barbara Ndambidi Yungi, Quentin Beaufort, Maxime Taverne, Nathalie Boddaert, Jean-François Mangin, Roman Hossein Khonsari, David Germanaud

## Abstract

Craniostenosis, a cranial malformation due to premature suture fusion, includes rare *FGFR*-related conditions such as Crouzon (CS), Muenke (MS), and Apert (AS) syndromes. These mutations affect skull but also brain development and are associated with variable neurodevelopmental outcomes. This brain impairment is still poorly described and understood, prompting for neuroimaging anatomical studies. We thus investigated regional brain growth and volumetric scaling in patients with these three syndromes and non-syndromic non-*FGFR*-related craniostenoses, using a normative modelling framework. We analyzed a rare set of brain MRI from 65 patients close to first cranial surgery time (30 CS among which 23 pre-surgery, 4 AS, 6 MS, 23 non-syndromic) and 130 typically developing controls. Millimetric 3D T1-weighted images were segmented into 6 cortical and 3 subcortical regions (AssemblyNet, Morphologist). After harmonization (ComBat), normative growth curves (Turner model) and static scaling curves (power-law model, effect of brain size regardless of age) were derived in controls. Patient deviations were assessed using permutation tests on mean normalized distance to typical model and abnormality rates (10th/90th percentiles). Patients with CS showed typical brain volumes for age except for enlarged ventricles, but slightly overscaled frontal volumes for brain size. Non-syndromic patients showed very similar volume distribution for age and brain size than CS ones. Patients with AS displayed increased total brain, temporal, cerebellar and central volumes for age, with overscaled temporal and cerebellar regions but underscaled frontal volumes for brain size. Patients with MS showed a similar, though milder, pattern: increased temporal volume for age, with temporal overscaling but frontal underscaling for frain size. Our findings reveal genotype-dependent brain anomalies in *FGFR*-related craniosynostoses, with a convergent pattern of temporal–frontal disproportions in AS and MS, associated with very specific mutations in the *FGFR2* and *3* genes respectively, contrasting with a much more typical brain organization in more heterogeneous CS and non-syndromic patients. Beyond the pathophysiological meaning of these results, which call for a larger similar study covering as wide a range of patient variability as possible, our work highlights the relevance of normative modelling approach for characterizing rare craniofacial disorders using clinical MRI datasets.

## 1 Introduction

Craniosynostoses are congenital craniofacial malformations resulting from the premature fusion of one or more cranial and/or facial sutures, leading to abnormal skull shape and potentially impaired brain development and growth (Yasaroglu & Demir, 2023). Some forms of craniosynostoses are associated with mutations in the fibroblast growth factor receptor (*FGFR*) gene group (Webster & Donoghue, 1997), such as Crouzon syndrome, Apert syndrome, and Muenke syndrome. Their prevalence varies, with Crouzon syndrome occurring in 1/50,000 to 1/60,000 live births (Helman et al., 2014), Apert syndrome in 1/65,000 (Breik et al., 2016), and Muenke syndrome being more common at 1/10,000 births (Sabatino et al., 2004). Besides these rare syndromic forms, non-syndromic craniosynostoses are much more common and occur in 5.2 of every 10,000 live births. In these conditions, the skull malformation occurs in isolation, with no identified mutation or evident extra-cranial phenotypic expression in most cases, particularly without known specific cerebral phenotype (Alperovich et al., 2025).

Syndromic craniosynostoses are characterized by extensive phenotypic features including cranial, facial, cerebral and extracephalic features. Each syndrome presents with distinctive cranial abnormalities. Crouzon syndrome is characterized by midface hypoplasia (Levesque et al., 2019) and is associated with variability in suture closure, with different sutures affected to different degrees depending on the patient (Azoury et al., 2017) leading to variable levels of skull growth anomalies (Delassus, 2025). Apert syndrome also features midface hypoplasia and is generally associated with bicoronal craniosynostosis (Giancotti et al., 2014). Muenke syndrome generally has a less severe facial involvement and is associated with bicoronal craniosynostosis in most cases (Mathijssen, 2015).

The cerebral phenotype of syndromic craniosynostoses is less clearly defined that the skeletal phenotype and varies in severity regarding brain structure and cognitive functions. Surgical interventions may influence outcomes (Mathijssen, 2015), but a cognitive and behavioral phenotype is frequently observed despite optimal management that may lead to severe neurodevelopmental impairment (Mathijssen, 2015). The functional phenotype in Crouzon syndrome is varied and imprecisely documented, visual impairment affecting up to 40% of patients (Bowling & Burstein, 2006), and hearing loss in 74% (Agochukwu et al., 2014). Intellectual impairment is reported in around 50% of patients with Apert syndrome (Mathijssen, 2015). In Muenke syndrome, intellectual disability occurs in around 35% of cases (Mathijssen, 2015). In addition, recurrent neurological abnormalities are associated with these conditions. In Crouzon syndrome, intracranial hypertension occurs in 40-63% of cases (Levesque et al., 2019), (Spruijt et al., 2016), as well as ventriculomegaly (38–70%) (Doerga et al., 2022), hydrocephalus (29%) (Coll et al., 2015), and Chiari I malformations (70%) (Cinalli et al., 2005). Apert syndrome is associated with ventriculomegaly in approximately 48.5% of cases (Breik et al., 2016), hydrocephalus in 9% (Giancotti et al., 2014), temporal cortical dysgyria (Breik et al., 2016, (Cohen & Kreiborg, 1990), abnormalities of the corpus callosum and limbic system (de León et al., 1987, Maksem & Roessmann, 1979), and generalized white matter reduction (Raybaud & Di Rocco, 2007). Muenke syndrome has been related to an increase in the volume of the posterior fossa and cerebellum (Rijken et al., 2015).

Nevertheless, although these syndromes differ clinically, they share a close genetic relationship through activating mutations affecting *FGFR* genes (*FGFR2* and *FGFR3*). These mutations notably lead to bone abnormalities and premature fusion of cranial sutures (Azoury et al., 2017). Both *FGFR2* and *FGFR3* alternatively splice their mRNA, producing isoforms that bind different fibroblast growth factors (FGFs) (Galzie et al., 1997), this ligand-binding specificity being crucial for embryonic development and organogenesis. Most *FGFR2* mutations occur in exons IIIa and IIIc, essential regions for receptor function (Chim et al., 2011), the FGFR2c isoform (alternative exon IIIc) being particularly expressed in the mesenchymal tissues forming the flat bones of the developing skull (Chim et al., 2011). A large diversity of pathogenic *FGFR2* variants is responsible for Crouzon syndrome while in Apert syndrome, p.S252W and p.P253R elective mutations alter the ligand affinity of FGFR2c (Chim et al., 2011), enabling abnormal binding to specific FGFs. In addition to osteogenesis, FGFs are also involved in brain development, neuronal maturation, neuroprotection and neurometabolic regulation. For example, FGF1 and FGF2 contribute to neuronal growth and plasticity, with FGF2 also stimulating osteogenesis (Reuss et al., 2003). Therefore, activating *FGFR* mutations is expected to affect both skull morphogenesis and brain development, underlying the intricate phenotypes observed in these syndromes.

Despite these insights, the cerebral phenotype associated with *FGFR*-related craniosynostoses remains poorly understood, and epidemiological and clinical data are rare. Neuroimaging research based on magnetic resonance imaging (MRI) has just begun to explore brain structure with more details. A recent study reported that patients with *FGFR* mutations had increased intracranial volume compared to controls, while displaying relatively reduced cortical surface area, particularly in the parietal and occipital lobes (Wilson et al., 2022). The existing literature, however, reveals contradictory results. Studies of intracranial and cerebral volumes in craniosynostosis have reported heterogeneous results, ranging from increased volumes in some syndromes (Dubois et al., 2008) to normal volumes in others (Junn et al., 2023). These divergences may be attributed to variations in the types of syndromes, sample sizes and imaging methodologies. Overall, these observations highlight important lacunae in our understanding of the brain patterns associated with *FGFR* mutations. Indeed, understanding the mechanisms of brain disruption in *FGFR*-related craniosynostosis syndromes presents several challenges. Firstly, it is essential to improve our knowledge and description of observable brain abnormalities associated with the different syndromes. Secondly, it is crucial to distinguish the congenital cerebral alterations from secondary complications linked to the progression of the disease or the side-effects of therapeutic wandering and surgical treatments. Finally, identifying correlations between genotype and brain phenotype remains an essential objective for advancing diagnosis and personalized management.

Today, tertiary expert centers are able to gather relatively large clinical series of genotyped and perisurgically-imaged patients, allowing the study of the neuroanatomical phenotype of these syndromes in greater depth, while carefully controlling for confounding factors. Object-based neuroimaging strategies relying on a limited set of robust regions of interest and anatomic parameters (volumes, surfaces), combined with normative analysis methods (Marquand, 2016), are expected to be sensitive enough to detect individual brain abnormalities associated with *FGFR2* or *3* pathogenic variants, such as global or focal abnormalities in brain growth (size-for-age), or even abnormalities in brain scaling (size distribution among regions) independent of a global sizing (Fraize et al. 2023; Kerdreux et al., 2025).

Using a clinical MRI dataset of patients with craniosynostosis, including patients with Crouzon, Apert and Muenke syndromes but also patients with non-syndromic presentations, imaged before or close to first surgical step, and a large group of typically developing controls, we segmented, quality-checked and harmonized 8 cerebral volumes of interest at the scale of large cerebral regions. Firstly, we established typical normograms on controls, based on growth (cross-sectional effect of age) and static scaling models (size effect independent from age). Secondly, we tested whether the patient groups showed global or regional deviations from the volumes expected for their age. In a third step, we regressed out the age parameter, under the assumption of z-score stability during growth, and tested whether the patient groups showed regional deviations from the expected volume for brain size.

## 2 Material and Methods

### 2.1 Participants

Participants in this study were 40 patients diagnosed with *FGFR2* or *FGFR3*-related syndromic craniosynostoses, aged from 1.5 to 17 years (see details in *Sup. Figure S1*). These included 30 patients with Crouzon syndrome, either before surgery or shortly after a first operative time (M= 5.78y, SD = 3.0, sex ratio = 1.6), 4 patients with Apert syndrome before surgery (M= 14.11yy, SD = 3.1, sex ratio = 1) and 6 patients with Muenke syndrome shortly after a first operative time, M= 3.2y, SD = 2.0, sex ratio = 1). Patients were retrospectively recruited from a monocentric clinical series at the tertiary reference center for craniosynostoses at Necker - Enfants malades Hospital (NEM) in Paris between 2009 and 2023. A reference population of 23 non-syndromic craniosynostoses patients was recruited shortly after a first operation from the same site. They ranged in age from 1.8 to 14 years (M= 6.61y, SD = 3.6, sex ratio = 1.9).

Typically developing (TD) controls included 130 subjects, from 1.5 to 18 years old (M = 12.2, SD = 3.4, sex ratio = 1.0), with no report of family history of neurological or psychiatric condition. A subgroup of 32 TD subjects (M = 10.4, SD = 4.4, sex ratio = 1.5) was matched to the craniosynostosis group for the MRI acquisition site and sequences. Another subgroup of 71 TD subjects (M=3.22, SD=1.4, sex ratio = 1.2) was included from the Baby Connectome Project (Howell et al., 2019). The remaining 27 TD subject (M=8.0, SD=2.33, sex ratio =1.1) were part of a previously published study (Bouyeure et al., 2021).

### 2.2 MRI Acquisitions

Five different 3DT1 T1-weighted UF-GRE (Ultra-Fast Gradient Echo) sequences were used for the acquisitions at the NEM hospital site. These included two sequences acquired at 3 Tesla (3T) and three sequences acquired at 1.5 Tesla (1.5T), on three different General Electric scanners (2 scanners at 1.5T and 1 scanner at 3T). Sequences were all at isometric or quasi-isometric millimeter resolution (in which case volumes were resampled to 1mm isotropic)as part of the flowchart process. The control group from the NEM site was acquired over the same period and with the same sequences, thus ensuring comparability between patients and controls within this site.

This study was conducted in accordance with the principles of the Declaration of Helsinki. Subjects’ data were studied in accordance with French regulation, following approval by the institutional review board and committee of the Necker – Enfants Malades University Hospital (study No. AP-HP - NCK-2025-E-045 TH). Controls data were used within the framework of the ethical authorizations of the primary studies.

### 2.3 Processing

T1-weighted brain MRI were segmented into 8 regions using the following pipeline combining BrainVISA (Rivière et al., 2009), VolBrain (Manjón et al., 2022) and AssemblyNet (Coupé et al., 2019). Morphologist2021 (BrainVISA) (Mangin et al., 2004) provided the inner cortical surface mesh and a one voxel-thick skeleton of the cortical sulci. Vol2Brain provided the deep grey matter nuclei, the brainstem and cerebellum. AssemblyNet provided a BrainCOLOR-based detailed volumetric parcellation of the cortex which was merged into 6 cortical regions: frontal, central, temporal, parietal, occipital, and insula. Two steps of regularization of these parcels were performed to remove improper cross boundary labelling (for instance across sulcal boundaries): a volume-based one (one sequence of erosion, sulcal skeleton exclusion, largest component selection, dilation) and a surface-based one after projection of the parcels on the inner cortical surface (one sequence of erosion-dilation). Deep gray matter and brainstem were merged into a ‘braincore’ (regularized by morphological closure). The six cortical surface parcels were ultimately back-projected into the brain volume and expanded within the hemispheres to the braincore boundaries (multilabel front propagation) to define 6 volumetric hemispheric ROIs. All the morphological operations were performed with the BrainVISA toolbox (see Figure 1 for an overview of the processing pipeline).

**Figure 1:**
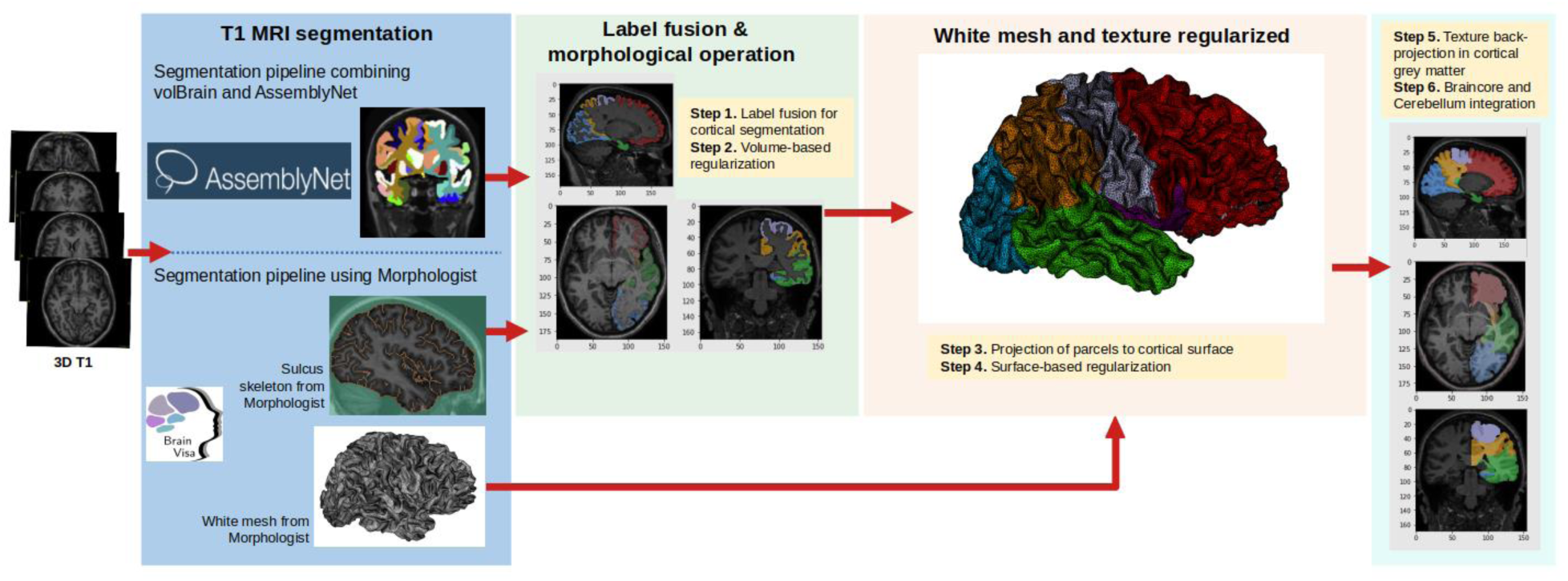
Processing pipeline for the segmentation of T1-weighted brain MRI into eight neuroanatomical regions of interest. The workflow combined VolBrain, AssemblyNet, and BrainVISA for brain structure segmentation and cortical parcellation. Regularization and merging steps refined the regions.

### 2.4 Statistical Analysis

All statistical analyses were carried out using Python and its packages pycombat (v0.20; (Behdenna et al., 2020)), scikit-learn (v1.2.2; (Pedregosa et al., 2011)) scipy (v1.11.4; (Virtanen et al., 2020)) and statsmodels (v0.14.0; (Seabold & Perktold, 2010)). We used the Benjamini and Yekutieli (Benjamini & Yekutieli, 2001) false discovery rate (FDR) control method to adjust p-values, to control the proportion of false positives among significant results in a context of multiple dependent comparisons.

#### 2.4.1 Adjusting for confounders factors or variables of noninterest

Although the NEM site included several sequences and scanners, it was treated as a single site for batch correction as patients and controls were acquired using the same panel of sequences. To ensure that this approach did not compromise the effectiveness of the harmonization and that no residual subgroup structure remained after harmonization, a post hoc analysis (*Sup. Table S1* for details) was carried out regarding the 32 control subjects from the NEM site. After harmonization of the whole control group, we made a 4-fold partition of the NEM control group (subsets of 8 subjects) and fitted 4 times the normative models for age including only 3 of the 4 NEM subsets into the whole control group. For each fold, a shuffle and split permutation test (see below) was performed to assess for the presence of a significant mean deviation for the normative model of age in the subset not included in the model fit. In none of the 4 folds did we did not observeddeviation in any of the 4 folds, testifying for a good correction of the site effect on the anatomical parameters studied, despite the heterogeneous nature of the NEM site.

The native volumes were adjusted using ComBat (Behdenna et al., 2020), for the effect of acquisition site and sex in order to perform normative analyses of the age effect and static scaling as proposed in previous studies (Fraize et al. 2023; Kerdreux et al., 2025). Sex was adjusted a priori to avoid dividing the study groups and to preserve maximum statistical power notably when establishing the reference curves. Combat was based on a linear model with empirical Bayesian estimation to harmonize batch effects (Behdenna et al., 2020). The site effect was corrected first, followed by the sex effect, while preserving age and group (patients, controls).

#### 2.4.2 Assessing the effect of craniosynostoses on age-related brain volume dynamics

The mean lateralization index (2x(L-R)/(L+R)) was never significantly different from 0.05 and we did not have any a priori assumptions about lateralization abnormalities in the patient population, so we grouped right and left volumes by equal structures.

We analyzed the effect of age on total brain volume (total parenchymal brain volume, TBV) and regional volumes using cross-sectional data. Normative growth curves were fitted to the control group using a Turner model (Equation 1) (Turner et al., 1976) providing a more flexible fit than the von Bertalanffy or Michaelis models thanks to an additional degree of freedom (Germanaud et al., 2012).

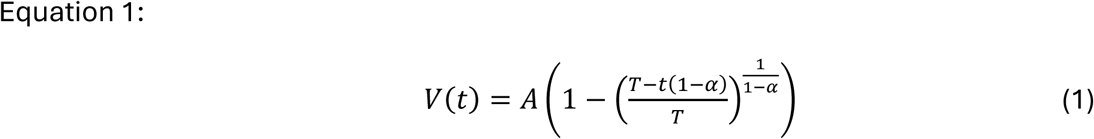

where *V* is the volume, *t* is the age, *A* is a volumetric constant parameter, *T* is a temporal constant, and *α* is a dimensionless exponent.

Note that the origin of growth time was set to the beginning of the 3rd trimester (-0.25 y) to ensure biological plausibility of the normative curves, in particular brain volumes at birth consistent with values reported in the literature (Lechpammer et al., 2021 ; Holland et al., 2014). Indeed, the Turner growth model does allow to model the inflection point occuring around 32–33 gestational weeks, so cannot be used from earlier on (i.e. origin cannot be set to the beginning of development).

The homoscedasticity of the residuals and the normality of the variance were verified in the control group to validate the conditions for using the following statistics and the z-scores both for visualization and to correct for the effect of age in the static scaling analysis (*Sup. Table S2* for details).

The mean deviation between groups and the reference model were assessed using a permutation test adapted to the situation of discrepancy between test and learning group (shuffle and split test (Germanaud et al., 2014)). To increase sensitivity in the event of non-homogeneous patient dispersion (test for differences in the distribution of percentiles between groups), patients were classified according to their position relative to the normative curves (below the 10th percentile, between the 10th and 50th percentiles, between the 50th and 90th percentiles, and above the 90th percentile) and a proportionality test was performed (Fisher’s exact 4-category test).

#### 2.4.3 Assessing the effect of craniosynostoses on static-scaling

For the static brain scaling analysis, the effect of age was adjusted by projecting patient volumes to a reference age of 18 years, while retaining their z-score. This age corresponded to the highest extremity of the cohort and to a point where the normative curve reaches its asymptote. This adjustment strategy is thus under the reductive assumption, yet frequently observed in clinical practice (Park et al., 1997; Wright et al., 2011), that z-scores remain stable throughout growth after the age of 18 months.

We then modelled the scaling relationship between the volume of the different brain regions and the TBV, using a non-linear power law. This model allows to characterize size-dependent proportional variations, away from a strict isometric (aka homothetic) relationship (Bookstein et al., 2006; de Jong et al., 2017; Germanaud et al., 2012; Liu et al., 2014).

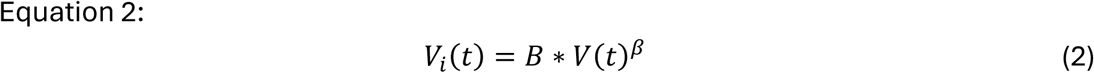

where *V_i_* is the volume of interest, *t* is the age, B and β (scaling exponent) are two constants fitted to the data.

This model (Equation 2) was fitted to the data from the control group. We assessed the homoscedasticity of the residuals then the effect of craniosynostoses on scaling by comparing the mean differences between groups using a shuffle and split test, and by testing whether there was an excess of patients above the 10th or 90th percentile of the reference distribution using a proportion test.

## 3 Results

The analyses were performed on 193 participants (40 patients diagnosed with syndromic craniosynostoses, 23 non-syndromic craniosynostoses patients and 130 TD subjects). Table 1 presents the clinical data of the patient groups.

**Table 1:** Clinical characteristics of the cohort.

|  | Crouzon group<br>n=30 | Apert group<br>n=4 | Muenke group<br>n=6 | Non-syndromic group<br>n=23 | Controls<br>n=130 |
|---|---|---|---|---|---|
| Age at MRI, mean in years (SD) | 5.78 (3.0) | 14.11(3.1) | 3.2 (2.0) | 6.61(3.6) | 12.2(3.4) |
| Sex ratio | 1.6 | 1 | 1 | 1,9 | 1 |
| Timing of MRI: preoperative/postoperative (average time after first surgery in years, in case) | 23/7 (1.55) | 4/0 | 0/6 (2.02) | 0/23 | 130 |
| Closure pattern: bicoronal synostosis/ pansynostosis/ sagittal synostosis/ without closed suture | 10 /12 /7 /1 | 4/0/0/0 | 6/0/0/0 | 7/4/12/0 | 0/0/0/130 |
| *FGFR* variants | 13 *FGFR2* variants \* | *FGFR2* p.Ser252Trp, *FGFR2* p.Pro253Arg | *FGFR3* p.Pro250Arg |  |  |

\*: p.Cys278Phe, p.Cys342Trp, p.Cys342Tyr, p.Cys278Phe, p.Gly338Glu, p.Pro361Pro, p.Ser347Cys, p.Ser354Cys, p.Thr320Ala, p.Trp290Gly, p.Tyr381Asn, p.Tyr105Cys, p.Val359Phe

We obtained normative curves based on harmonized brain volumes derived from a series of control subjects. These curves (Figure 2) represented the expected distribution of volumetric measurements across the population and served as a reference for comparing patient data.

**Figure 2:**
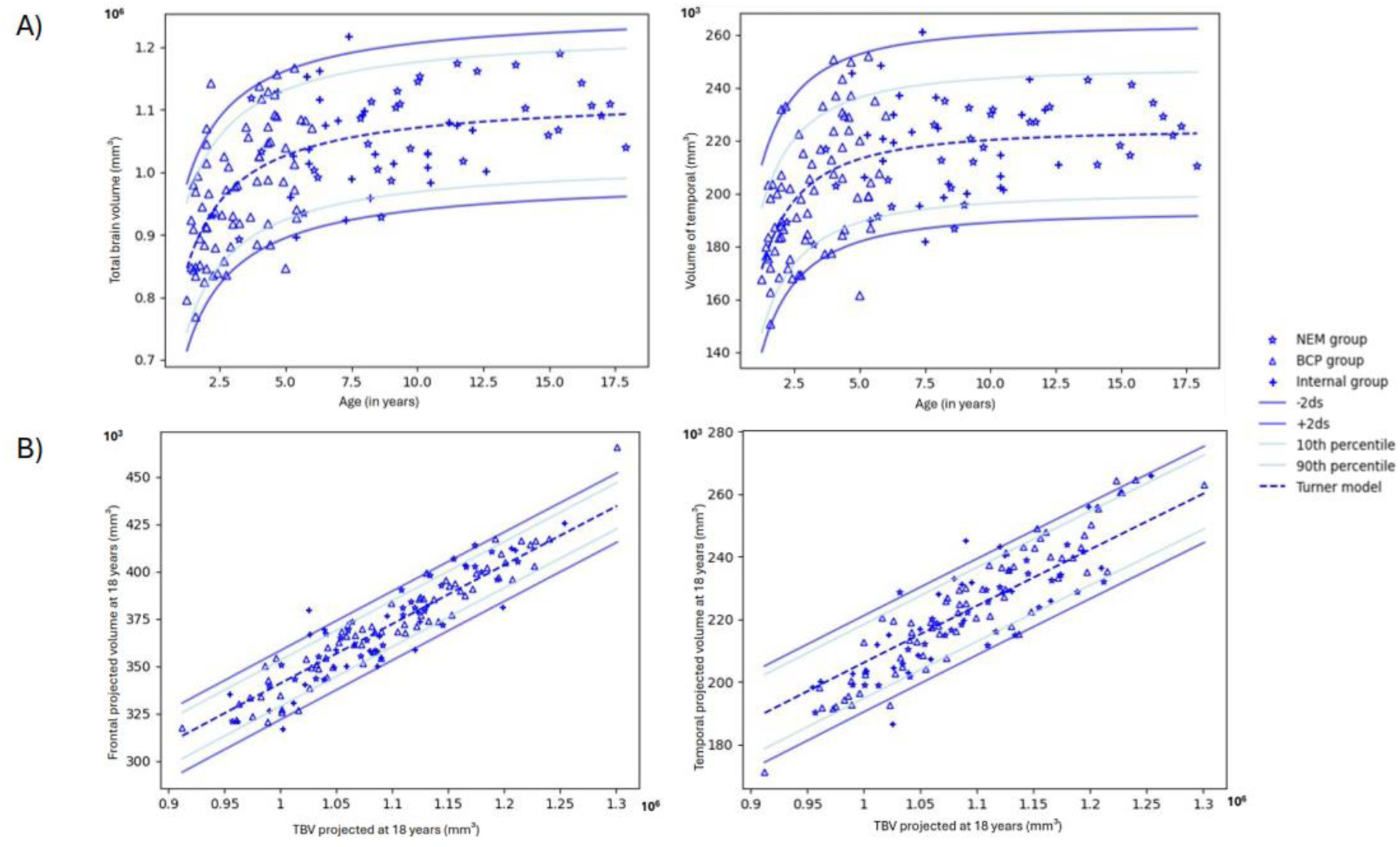
Normative volumetric charts derived from harmonized control data. (A) Cross-sectional growth curves of the total brain volume (TBV) and temporal lobe. (B) Static scaling curves showing the temporal lobe volume and the frontal lobe volume, each projected at 18 years, in relation to residual TBV projected at 18 years. Controls are represented with different symbols according to their origin: stars for the NEM group (*), triangles for the BCP group (Δ), and crosses for the internal group (+). The blue curves represent the 10^th^ to 90^th^ percentiles and ±2 standard deviations of the normative distribution. Full normative curves for all cerebral regions are available in the supplementary material (*Sup. Figure S2*).

### 3.1 Normative growth analyses (cross-sectional age-effect)

Subjects with Crouzon syndrome presented volumes (TBV and regional brain) similar to those of the control group (p-corrected>0.05) except for increased lateral ventricles volumes (p<10^−8^). In contrast, the volumes of the Apert syndrome group were larger than those of the control group for the central (p<10^−8^), parietal (p<0.005), temporal (p<10^−8^), lateral ventricles (p<10^−8^), cerebellum (p<10^−8^) and TBV (p<10^−3^) volumes (Figure 3, Table 2). The temporal volume of patients with Muenke syndrome was larger than controls for age (p<10^−8^). The volumes of non-syndromic patients were comparable to controls (Figure 3, Table 2).

**Figure 3:**
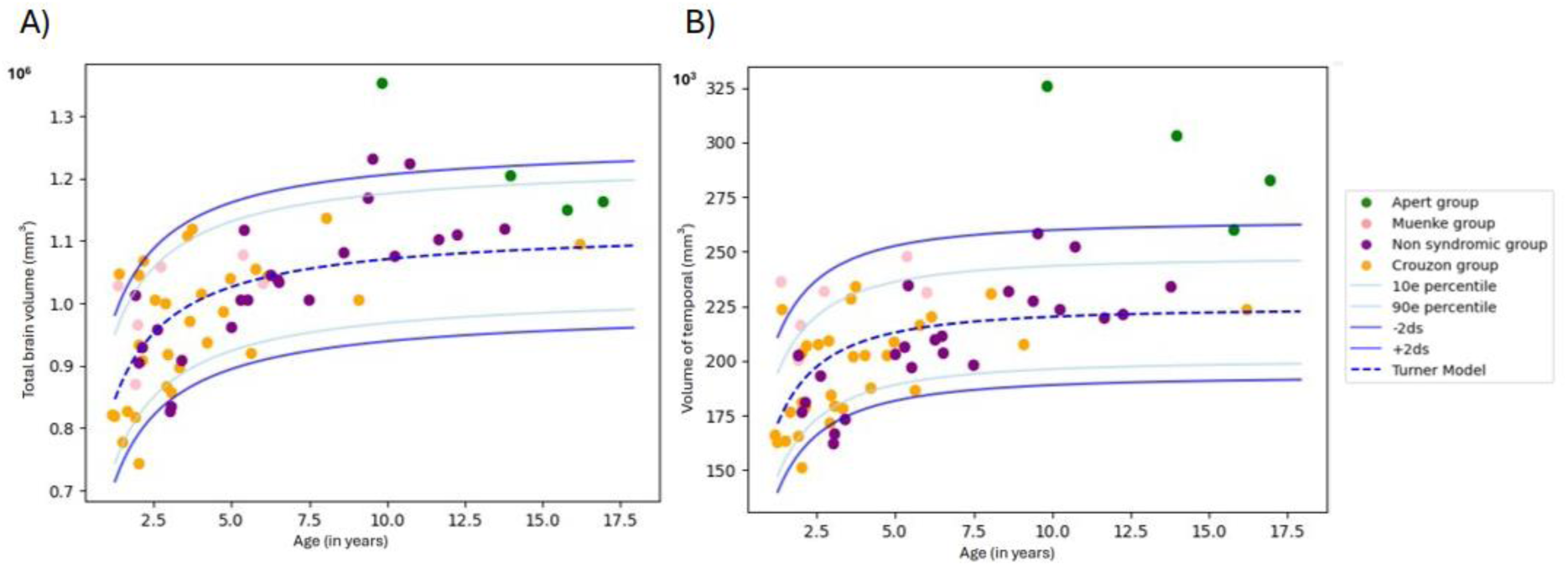
Age effect charts for total brain volume (TBV) (A) and temporal lobe volume (B) in the patient groups (colored dots: green for Apert syndrome, pink for Muenke syndrome, purple for non-syndromic, orange for Crouzon syndrome) compared with the normative curves for the control group. The fitted model curve for controls is shown as a blue dotted line. The other blue lines represent the 10^th^ and 90^th^ percentiles, as well as ±2 standard deviations of the normative distribution. Charts for other regional volumes are provided in the supplementary material (*Sup. Figure S2*).

**Figure 4:**
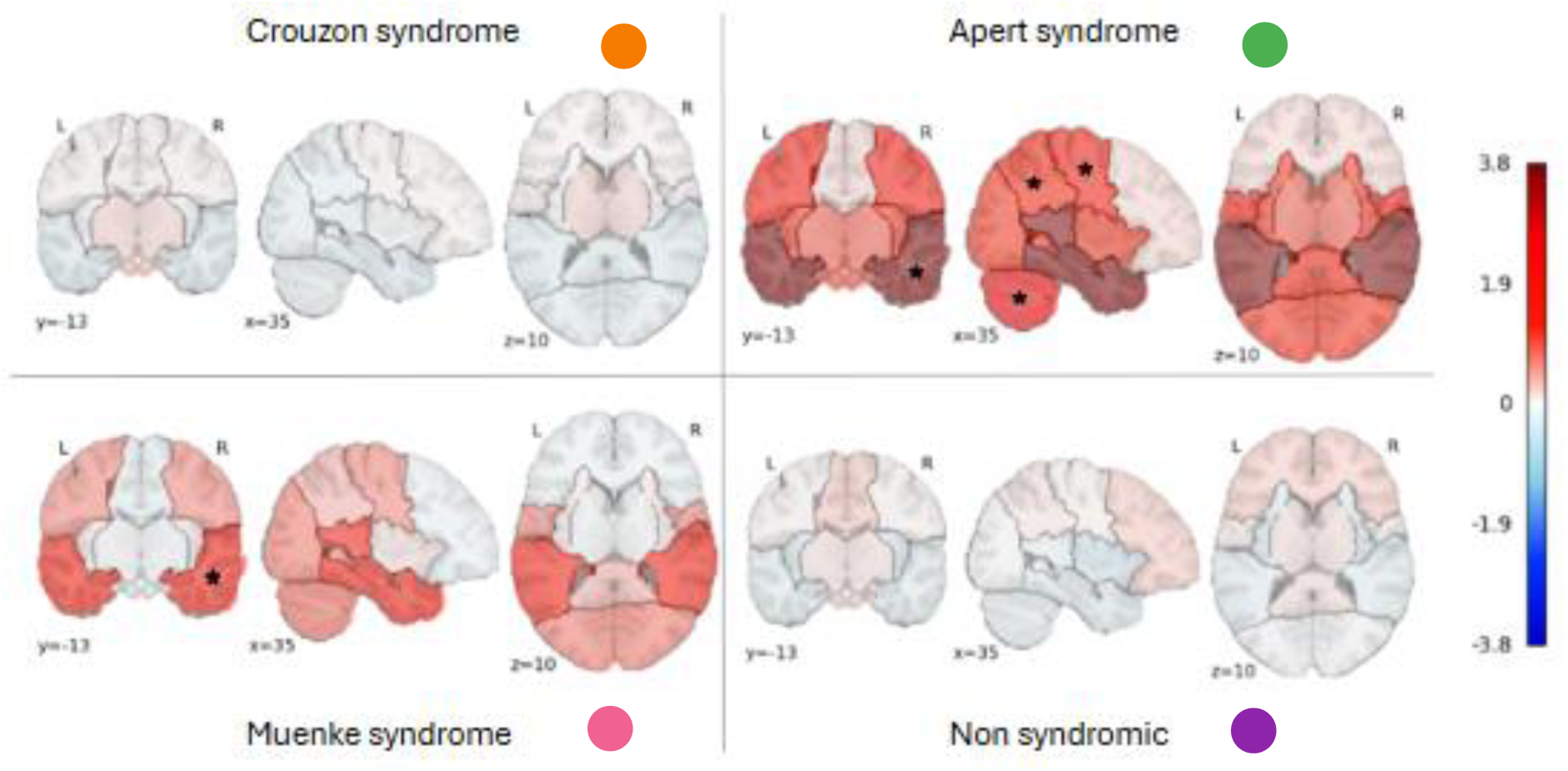
Pattern and extent of the difference between patients and controls for age effect. Maps represent the mean z-score difference between each patient group and the control group for the brain regions, showing deviations from the expected volume for age. The colormap encodes the intensity of the deviation. Asterisks (**★**) indicate regions where the deviation is statistically significant (p < 0.05, permutation test). Colored dots identifying patient groups (green for Apert, pink for Muenke, orange for Crouzon, purple for non-syndromic) follow the same color codes as in the clinical summary table. Ventricles were not included in this figure.

**Table 2:**
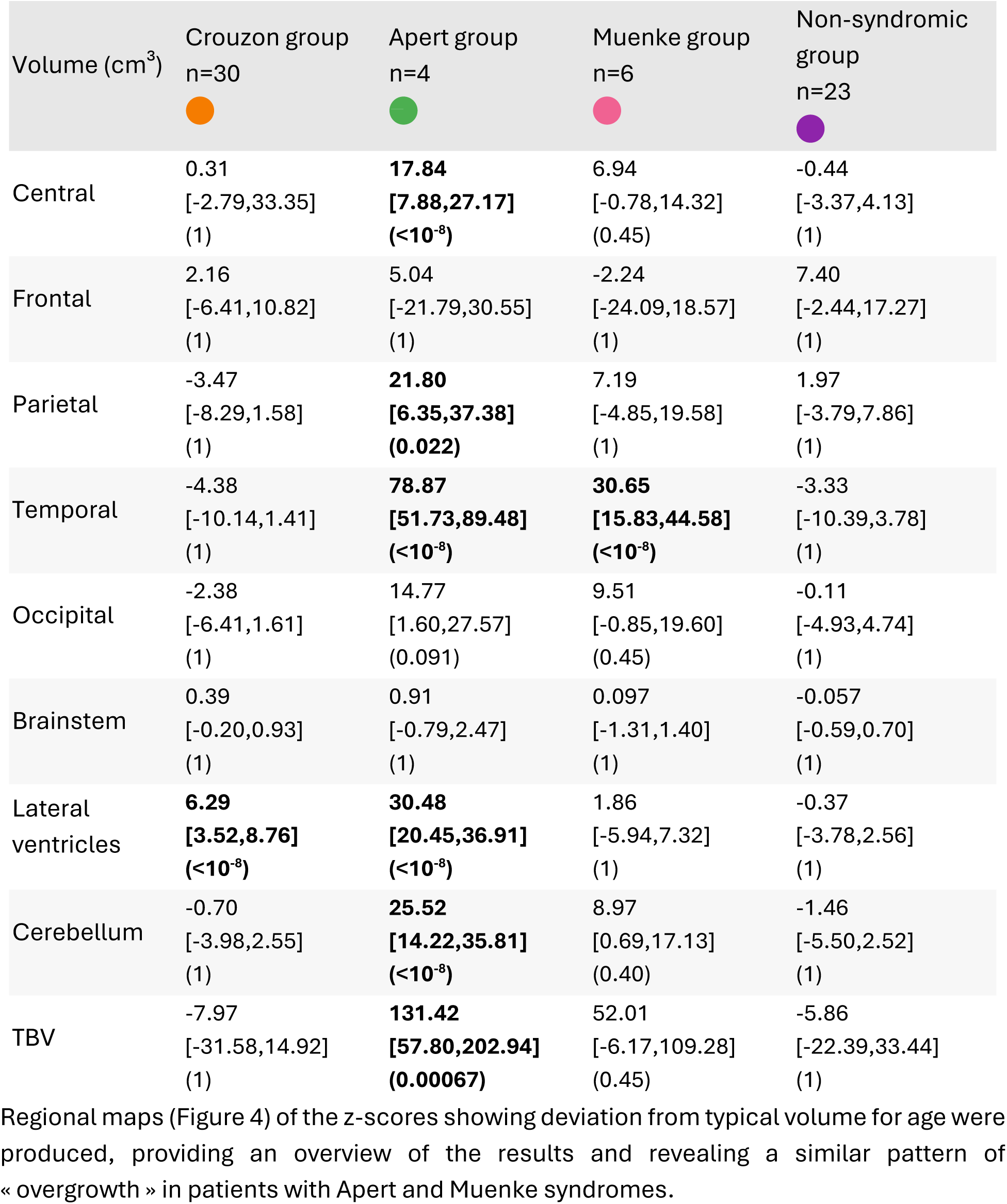
Tests evaluating the mean deviations of the age effect from controls. Values are shown as mean residual difference [95% confidence interval], with p adjusted using the FDR-BY method. Positive values indicate volumes larger than expected for age; negative values indicate volumes smaller than expected. Abbreviations: TBV = Total Brain Volume.

| Volume (cm<sup>3</sup>) | Crouzon group<br>n=30 | Apert group<br>n=4 | Muenke group<br>n=6 | Non-syndromic group<br>n=23 |
|---|---|---|---|---|
| Central | 0.31<br>[-2.79,33.35]<br>(1) | **17.84**<br>**[7.88,27.17]**<br>**(<10<sup>-8</sup>)** | 6.94<br>[-0.78,14.32]<br>(0.45) | -0.44<br>[-3.37,4.13]<br>(1) |
| Frontal | 2.16<br>[-6.41,10.82]<br>(1) | 5.04<br>[-21.79,30.55]<br>(1) | -2.24<br>[-24.09,18.57]<br>(1) | 7.40<br>[-2.44,17.27]<br>(1) |
| Parietal | -3.47<br>[-8.29,1.58]<br>(1) | **21.80**<br>**[6.35,37.38]**<br>**(0.022)** | 7.19<br>[-4.85,19.58]<br>(1) | 1.97<br>[-3.79,7.86]<br>(1) |
| Temporal | -4.38<br>[-10.14,1.41]<br>(1) | **78.87**<br>**[51.73,89.48]**<br>**(<10<sup>-8</sup>)** | **30.65**<br>**[15.83,44.58]**<br>**(<10<sup>-8</sup>)** | -3.33<br>[-10.39,3.78]<br>(1) |
| Occipital | -2.38<br>[-6.41,1.61]<br>(1) | 14.77<br>[1.60,27.57]<br>(0.091) | 9.51<br>[-0.85,19.60]<br>(0.45) | -0.11<br>[-4.93,4.74]<br>(1) |
| Brainstem | 0.39<br>[-0.20,0.93]<br>(1) | 0.91<br>[-0.79,2.47]<br>(1) | 0.097<br>[-1.31,1.40]<br>(1) | -0.057<br>[-0.59,0.70]<br>(1) |
| Lateral ventricles | **6.29**<br>**[3.52,8.76]**<br>**(<10<sup>-8</sup>)** | **30.48**<br>**[20.45,36.91]**<br>**(<10<sup>-8</sup>)** | 1.86<br>[-5.94,7.32]<br>(1) | -0.37<br>[-3.78,2.56]<br>(1) |
| Cerebellum | -0.70<br>[-3.98,2.55]<br>(1) | **25.52**<br>**[14.22,35.81]**<br>**(<10<sup>-8</sup>)** | 8.97<br>[0.69,17.13]<br>(0.40) | -1.46<br>[-5.50,2.52]<br>(1) |
| TBV | -7.97<br>[-31.58,14.92]<br>(1) | **131.42**<br>**[57.80,202.94]**<br>**(0.00067)** | 52.01<br>[-6.17,109.28]<br>(0.45) | -5.86<br>[-22.39,33.44]<br>(1) |

Regional maps (Figure 4) of the z-scores showing deviation from typical volume for age were produced, providing an overview of the results and revealing a similar pattern of « overgrowth » in patients with Apert and Muenke syndromes.

The count of subjects with abnormal values (>90^th^ <10^th^ percentiles of the reference curve) revealed very similar patterns of abnormalities. The detailed comparison testing is presented in supplementary material (*Sup. Table* S3).

### 3.2 Normative scaling analyses (brain volume distribution for size)

In the Crouzon syndrome group, the lateral ventricles were excessively large for the TBV (or oversized) (p<10^−8^) but conversely, the frontal lobe was undersized (p=0.022). The patients with Apert syndrome presented with oversized temporal lobes and lateral ventricles (p<10^−8^ for both) but undersized frontal lobes and cerebellum (p<10^−8^ and p=0.0044 respectively). In the Muenke syndrome group, the frontal lobes were undersized and the temporal was oversized (p<10^−8^ for both). In the non-syndromic group, the frontal lobes were oversized (p=0.029) and the temporal was undersized (p=0.029) (Figure 5, Table 3). The count of subjects with abnormal values (>90^th^ <10^th^ percentile of the reference curve) identified and located very similar abnormalities. Scaling charts and full test results can be found in supplementary material (*Sup. Table* S4 and *Sup. Figure* S3). Regional maps of the z-scores showing deviation from typical volume for brain size were produced, giving an overview of the results and revealing a similar pattern of disproportion in patients with Apert and Muenke syndrome on the one side, patients with Crouzon syndrome and non-syndromic craniosynostoses on the other.

**Figure 5:**
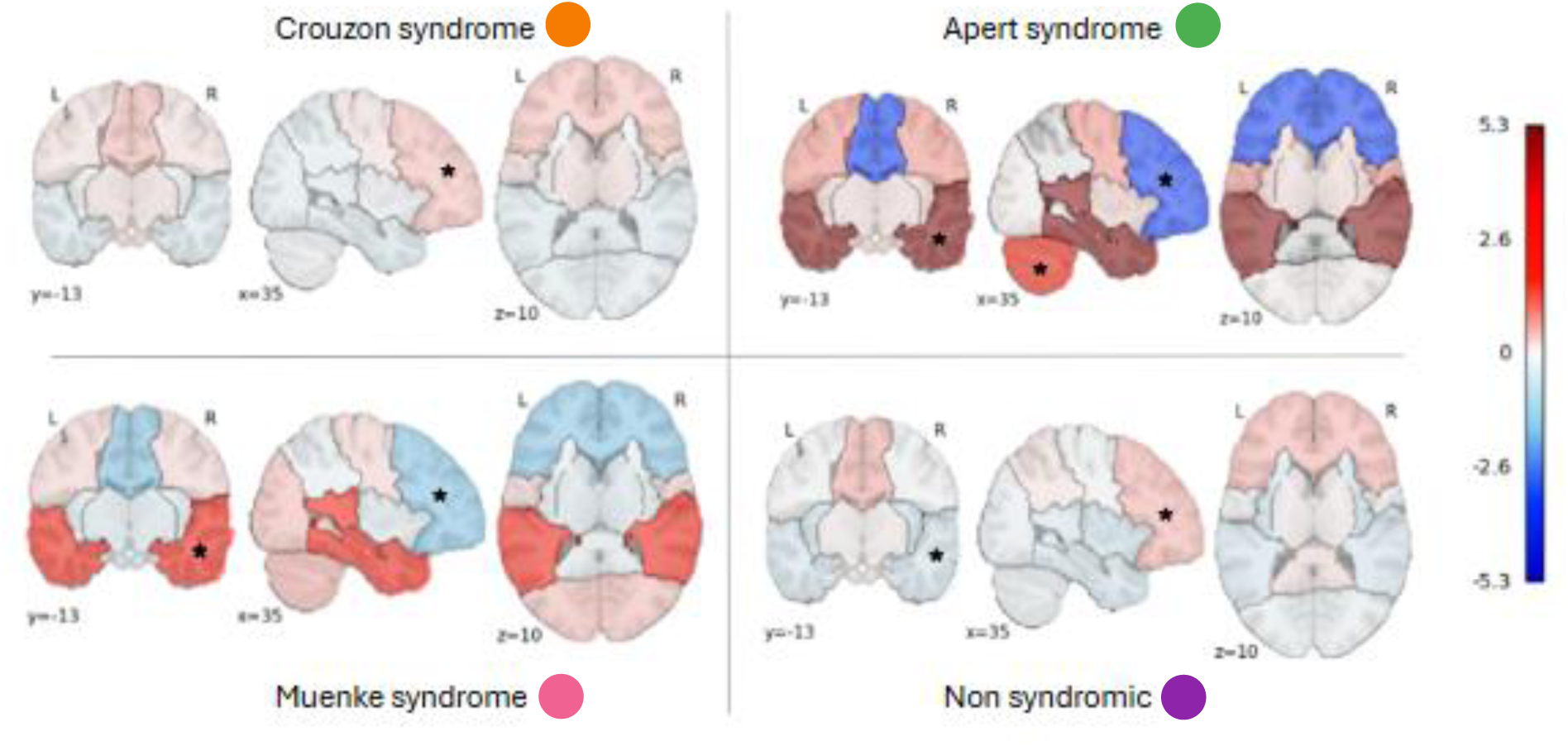
Pattern and extent of the difference between patients and controls in static scaling. Maps represent the mean z-score difference between each patient group and the control group for the brain regions, showing deviations from the expected regional volume for a given total brain volume (TBV). The colormap encodes the intensity of the deviation. Asterisks (**★**) indicate regions where the deviation is statistically significant (p < 0.05, permutation test). Colored dots identifying patient groups (green for Apert, pink for Muenke, orange for Crouzon, purple for non-syndromic) follow the same color codes as in the clinical summary table. Ventricles were not included in this figure.

**Table 3:** Tests evaluating the mean deviations from control in static scaling. Mean residual differences in regional brain volumes (cm³) projected at 18 years of age between each patient group and the control group, after adjusting for total brain volume (TBV) projected at 18 years of age. Values are shown as mean residual difference [95% confidence interval], with p adjusted using the FDR-BY method. Positive values indicate volumes oversized relative to total brain size; negative values indicate undersized volumes.

| Volume (cm<sup>3</sup>) | Crouzon group<br>n=30 | Apert group<br>n=4 | Muenke group<br>n=6 | Non-syndromic group, n=23 |
|---|---|---|---|---|
| Central | 0.99<br>[-0.95,2.93]<br>(1) | 4.92<br>[-1.24,10.54]<br>(0.52) | 1.30<br>[-3.70,5.84]<br>(1) | -0.12<br>[-2.54,2.13]<br>(1) |
| Frontal | **4.86<br>[1.72,7.96]<br>(0.022)** | **-39.06<br>[-49.30,-28.97]<br>(<10<sup>-8</sup>)** | **-19.74<br>[ -27.74,-11.58]<br>(<10<sup>-8</sup>)** | **5.41<br>[1.65,9.14]<br>(0.029)** |
| Parietal | -1.98<br>[-4.59,0.60]<br>(1) | -1.84<br>[-9.57,5.81]<br>(1) | 2.12<br>[-8.54,3.97]<br>(1) | 0.92<br>[-2.20,3.93]<br>(1) |
| Temporal | -2.53<br>[-5.11,0.21]<br>(0.78) | **41.58<br>[33.75,49.59]<br>(<10<sup>-8</sup>)** | **19.19<br>[12.79,25.80]<br>(<10<sup>-8</sup>)** | **-4.65<br>[-7.71,-1.44]<br>(0.029)** |
| Occipital | -1.29<br>[-3.59,1.13]<br>(1) | -3.52<br>[-10.63,4.17]<br>(1) | 2.10<br>[-3.92,8.43]<br>(1) | -0.94<br>[-3.75,1.92]<br>(1) |
| Brainstem | 0.31<br>[-0.23,0.82]<br>(1) | -0.30<br>[-1.41,1.81]<br>(1) | -0.59<br>[-1.97,0.60]<br>(1) | -0.047<br>[-0.58,0.65]<br>(1) |
| Lateral ventricles | **7.23<br>[4.39,9.73]<br>(<10<sup>-8</sup>)** | **24.38<br>[14.97,30.77]<br>(<10<sup>-8</sup>)** | 1.45<br>[-5.76,6.85]<br>(1) | -0.73<br>[-4.09,2.18]<br>(1) |
| Cerebellum | -0.48<br>[-3.67,2.68]<br>(1) | **19.71<br>[9.75,29.16]<br>(0.0044)** | 5.91<br>[-1.70,13.47]<br>(1) | -1.70<br>[-5.45,2.09]<br>(1) |

## 4 Discussion

Our study proposed a normative approach based on a rare set of clinical MRI to analyze cerebral volumetric regional profiles in syndromic craniosynostosis linked to *FGFR2* and *FGFR3* mutations. Our results highlighted syndrome specific cerebral alterations, particularly a contrasting pattern of growth and sizing (static-scaling) anomalies common to Apert and Muenke syndromes, albeit with a smaller effect size for the latter. Conversely, both patients with Crouzon syndrome and non-syndromic patients showed brain volume distributions close to typical individuals, with the mild singularities identified being similar in both conditions. In addition to a contribution to a better understanding of craniosynostosis linked to *FGFR* variants, our results also illustrate the potential of clinical data inscribed into a methodological framework exploiting the current reference datasets and harmonization tools.

### 4.1 Small groups but rather contrasted syndromic effects

The results indicated that Apert syndrome was associated with significant global and regional volumetric abnormalities. Apart from an enlargement of the lateral ventricles, the total brain parenchyma was overgrown so as the temporal lobes, consistent with some rare reports from the literature (increased intracranial volume (Wilson et al., 2022), temporal lobe abnormalities (Raybaud & Di Rocco, 2007)). Notably, this overgrowth also concerned other brain regions, such as particularly the cerebellum, but spared the frontal lobes. These differences were ultimately reflected in an atypical distribution of brain volume for brain size, with oversized temporal and cerebellar regions and relatively undersized frontal regions (for a brain of this size). As such, it may be interesting to look at this uneven distribution from another point of view, taking the frontal lobe - of expected size for age - as a reference, which highlights a gradient of oversizing for the rest of the cerebral regions in Apert syndrome, of which the temporal and, to a lesser extent, the cerebellar regions constitute the extrema (see this representation in Supplementary Material, *Sup. Figure S5*). Indeed, the very small number of patients with Apert syndrome, (n=4) constituting more a small case series than a representative group of the Apert syndrome population, tends to weaken these results. Their significance was supported by large effect sizes, but a selection bias is difficult to rule out. However, it should be remembered that the only active selection made was on the quality of the MRI scans and their compatibility with the proposed computational treatments. With this in mind, the intra-group consistency of the anomalies and their concordance with the literature reinforced their reliability.

More unexpectedly, as suggested by no preliminary data, our analyses revealed a rather similar, albeit less intense, pattern of volumetric anomalies in Muenke syndrome, with temporal « overgrowth » and oversizing, associated with frontal undersizing for brain size. Here again, the small size of the group (n=6) makes the result prone to an even « random » selection bias, but the consistency over the 6 subjects seems an element of conviction. Interestingly, in a context where we could not detect any global « overgrowth » of the brain, the static-scaling analysis revealed an atypical pattern of volume distribution similar to the one observed in Apert syndrome.

By contrast, patients with Crouzon syndrome showed only ventricular abnormalities, with almost no significant cortical or subcortical volumetric alterations for age and a mild uneven distribution of brain volumes for brain size, a configuration eventually very similar to the one observed in non-syndromic craniosynostoses. The size of these two groups might suggest a lower risk of insufficient sampling of the population, Yet, given the heterogeneity at least of Crouzon syndrome compared with Apert and Muenke syndromes both genetically (large diversity of *FGFR2* variants in Crouzon syndrome, virtually unknown determinism in non-syndromic craniosynostosis) and phenotypically (among other things, diversity of sutural anomalies) – we could have under-sampled subgroups and masked their specific effects. Qualitative observation of the data or distribution tests did not support this hypothesis, which would nevertheless require larger numbers to be effectively tested. Furthermore, complementary analysis of the Crouzon syndrome and non-syndromic groups restricted to patients with bicoronal anomalies did not support an effect of sutural phenotype on cerebral parameters in our patients. Yet this pattern of regional brain growth and scaling rather similar to typically developing controls in patients with Crouzon syndrome might look surprising given the high frequency of neurological symptoms reported in this condition, such as raised intracranial pressure (Spruijt et al., 2016), Chiari (Cinalli et al., 2005) or neurodevelopment impairments (Wenger et al., 2020, Wilson, 2020). The ventricle enlargement was almost expected and of course limited in amplitude by the fact that we excluded the patient with more than mild ventriculomegaly. The mild frontal oversizing for brain size common with non-syndromic forms – probably at the expense of the temporal-occipital regions according to the pattern trending in the two average z-score maps – might be more informative and an avenue for further studies, though it could only be the residual of an abusive averaging between more contrasted subgroup effects.

### 4.2 Relevance of a genotype-dependent brain phenotype

From a clinical perspective, it makes sense that we found the main growth and sizing regional anomalies in the brain of the patients with the two syndromes more associated with neurodevelopmental disorders (50% of intellectual development disorder in Apert syndrome, 40% in Muenke syndrome with 20% epilepsy (Kruszka et al., 2016). In the more heterogeneous Crouzon syndrome, it is considered difficult to dissociate the mild cognitive and behavioral impairments from the complications associated to prolonged increased intracranial pressure, the variations in therapeutic sequences or the psychosocial repercussions (Rostamzad et al., 2024). In this respect, it seems consistent that the mild pattern of cerebral missizing observed in Crouzon syndrome was identical to that of non-syndromic patients, who typically have few associated neurodevelopmental disorders.

Yet, recent investigations in the neuropsychological profiles of patients with non-syndromic craniosynostoses seem to show that mild cognitive alterations are more prevalent than in control groups, independently from surgical management and thus potentially caused by intrinsic brain anomalies that may not be detectable by current imaging protocols (Olsson et al., 2023). In addition, even though few studies are available to assess the cognitive phenotype in Crouzon syndrome (Fernandes et al., 2016), neuropsychological signs in this syndrome seem to be highly variable with no current valid genotype-phenotype correlation. Here we excluded Crouzon syndrome patients with severe initial presentations such as cloverleaf skulls, where brain malformations may be present. For instance, a neuropathological investigation of fetuses with *FGFR2*-related syndromic craniosynostoses diagnosed as Pfeiffer syndrome (now considered as a variant of Crouzon syndrome) have reported temporal lobe malformations that reminded the malformations observed in *FGFR3*-related thanatophoric dysplasia, a lethal form of chondrodysplasia (Hevner et al., 2005, Khonsari et al., 2012). The absence of brain phenotype we report may thus be related to a selection bias. Nevertheless, extremely severe presentations such as cloverleaf skulls are not the most common clinical situation in this syndrome and our results, both for Crouzon syndrome and non-syndromic craniosynostoses, are in line with the fact that these two conditions have generally milder cognitive alterations than Apert and Muenke syndrome patients.

Genetically speaking, the converging pattern observed in Apert and Muenke syndromes, if confirmed on a larger number of subjects, is both unexpected and questioning from a mechanistic point of view, given that these syndromes are linked to very specific variants but in two different genes of the *FGFR* family. Although it is not possible to consider this type of correlation in our dataset, it is now possible to test whether greater deviations from typical brain growth or regional dimensioning is associated with greater severity of the neurodevelopmental disorders associated with these two syndromes. Conversely, the apparent consistency of the absence of any major neuroanatomical abnormality in the Crouzon syndrome spectrum shall not hinder from questioning the relevance of this finding. Indeed, the high heterogeneity of genetic variants in Crouzon syndrome compared with syndromes such as Apert syndrome (2 causal variants) or Muenke syndrome (only one) could account for the absence of a homogeneous trend towards regionalized brain volumetric dysgenesis. It is also possible that all the *FGFR2* variants involved in Crouzon syndrome share a relative lack of effect on brain growth (which would not exclude them having common or specific effects on other aspects of brain development or maturation), compared to the two Apert syndrome variants for instance.

To better understand this point, the spectrum of Crouzon syndrome shall probably described regarding the cognitive phenotype and its correlations with neuroanatomy and genotype. As mentioned above, the most severe variants of Crouzon syndrome in our cohort have been excluded from this study. Crouzon syndrome patients with cloverleaf skull and/or severe and early hydrocephalus do not have a specific genetic background reported to date. Despite this current limitation in the characterization of the cohort, a stratification of Crouzon syndrome patients according to cognitive status could still contribute to tackle an important question raised by our study. In fact, the brain malformations we report in Apert and Muenke syndromes could be: (1) common to all patients with FGFR mutations and severe brain involvement, including the excluded forms of severe Crouzon syndrome patients, or (2) relate to specific, still undescribed, mechanisms common to Muenke and Apert syndromes but not present in Crouzon syndrome. According to the current, very limited, available date in the literature, the first hypothesis seems the most relevant. In fact, as mentioned above, severe prenatal forms of FGFR2-related Crouzon syndrome (labeled as Pfeiffer syndrome) have been associated with major brain malformations resembling malformation in other conditions due to activating FGFR mutations such as FGFR3-related thanathophoric dysplasia. Furthermore, the very uncommon subtype of Crouzon syndrome due to FGFR3 mutations (Crouzon with acanthosis nigricans) seems to be associated with specific clinical severity and brain anomalies (Cornille et al., 2022, de Planque et al., 2021). These phenotypic overlaps in conditions due to different mutations may be related to the expression overlap of *FGFR* genes in the craniofacial region, both in humans and in mouse models. In brief, as there is no current argument to relate the mechanisms underlying Apert and Muenke syndromes, the proximity of their brain phenotype may correspond to the characterization of the severe side of a common spectrum of brain anomalies due to all activating FGFR mutations (Delezoide et al., 1998, Paek et al., 2009).

Furthermore, even though the genetics causes of the vast majority of non-syndromic craniosynostosis cases are still to be uncovered, several mutations have been associated with specific risks of cognitive impairment and potential brain anomalies (Timberlake et al., 2022). The non-syndromic group also shall benefit from more precise neuroanatomical characterization in relation with functional severity before being considered as a control group where the sole source of anomalies is the skull deformation.

Finally, our results also suggest that the genotype has a predominant influence on cerebral organization in craniosynostoses compared with the sutural phenotype, which could have played a direct role (by simple deformation or even developmental stress), or else be an indirect marker of cerebral dysgenesis. Indeed, even when restricting the analysis to patients with bicoronal synostosis, no convergence in volumetric pattern emerged between the Crouzon syndrome group (and the non-syndromic patients) and the Apert syndrome or Muenke syndrome groups. Also, the post-hoc analysis performed on the Crouzon syndrome patients did not show any major differences between the Crouzon syndrome subgroups according to the suture affected.

### 4.3 Interest, means and limits of normative analyses in clinical datasets

Neuroimaging research into rare conditions such as syndromic craniosynostosis is complicated by the lack of access to sufficient quality data in sufficient quantity. Our approach has been to combine a number of methodological tools implemented in recent years for neuroanatomical analysis in computational imaging, in order to make the most of the rare retrospective data set at our disposal. The main strategy had been to generate normograms reflecting the effect of age (cross-sectional) and size (static-scaling) in the context of typical development, which would be accurate enough and compatible with our patient data. To achieve this, we combined data harmonization tools, whose effectiveness in computational neuroanatomy has been widely documented, with explicit modeling of normative curves by rather simple biomathematical functions (few parameters to fit), which are relatively data-saving.

In terms of harmonizing data by correcting for batch effects, the main weakness of our approach lay in the heterogeneous nature of the patient acquisition site (NEM site). Indeed, even though the same panel of 3DT1 sequences was used for patients and controls from the NEM site, it was not possible to divide this site into as many sequences for the sake of parametrization. This is why we sought to verify whether this choice did not bias the analysis of mean divergence from the normative model obtained on all controls by a post-hoc test based on a partition of controls from the NEM site: none of the four partitions of eight controls considered as a group of interest (pseudo-patient) showed significant deviation from the norm for any of the anatomical metrics studied, validating the hypothesis that, within the precision limits of the study, our simplification of the batch effect model remained acceptable.

More generally, it would have been preferable to have more homogeneous data in terms of 3DT1 sequence parameterization, over and above the multisite character, even though the large neuroanatomical objects we studied are probably little sensitive to sequence effects due to the dilution of the small differences induced in the total volume of the object (unlike cortical objects, for example).Despite our reassuring post-hoc verifications, taking into account the sequence effect through the site effect is probably not optimal, but it is important to bear in mind the size of the main effects finally observed, particularly in the Apert and Muenke syndromes group, which it is hardly debatable and exceeds the magnitude of the noise or even the residual biases linked to the production of the normograms.

Regarding the modelling of age and size effects, the power law choice for scaling analysis rather consensual and its use was highly documented, including from our previous work. Modeling age effect forms a 130 subject large control group over a 16 years window with a rather simple (parsimonious parametrization) function is a less well-documented task. Interestingly, classical growth curves such as von Bertalanffy didn’t show convincing fits, with nearly asymptotic values reached as early as 4 to 5 years of age, which was not consistent with the literature (Peterson et al., 2018). We chose to rely on a more general model published by Turner and providing sufficient degree of freedom to fit a more convincing “growth” dynamic from our data set at the extent of only 3 parameters to fit. Eventually, within the limits of the available power, we found the residual from our models to show no heteroscedasticity, allowing for rather easy estimate of z-score and proper uses of the proposed statistics.

### 4.4 Other built-in limitations

Concerns about the sample size, particularly for Apert and Muenke syndromes, and about the setting-up of normative charts have been already mentionned but two important built-in limitations have still to be taken into.

First, we wanted to study the brains of patients with the least impact on development from mechanical complications such as hydrocephalus, or from the often-cumbersome therapeutic sequences, which also degrades the quality of brain images. For the sake of recruitment, we did include data from MRI performed before surgery, but also just in the aftermath of the first surgical step (may the only). This pragmatic and developmentally relevant choice nevertheless reduces the representativeness of the data, associated with the de facto exclusion of the large number of children who required early surgery, at an age when only T2 weighted images were available, which is often not usable (clinical 2D imaging) and in any case requires very different analysis strategies. Besides, potential effect of the first step of surgical procedures cannot be totally ruled out, even if a comparison between pre- and post-operative Crouzon syndrome (see supplementary material *Sup. Table S5*) revealed no significant difference, suggesting a limited effect in the short term, but a longitudinal study would be needed to validate this point.

Second, as in previous normative studies with medium sized control group (150-250), we favored statistical power by fitting the reference in the overall typically developing sample, while controlling for the effect of sex with data harmonization using ComBat (Fraize et al., 2023; Kerdreux et al. 2025). Indeed, many studies have shown that total brain volume is generally higher for men even if these differences were more limited at regional level, particularly when volumes were corrected for total brain size (TBV) (Mills et al., 2016). The exploration of any interactions at the brain level between sex and *FGFR*-related craniosynostosis, that definitely have been described in many developmental conditions (Postic et al., 2024, Smith et al., 2014), was then not considered. Provided larger number of subjects, analyses stratified by sex should certainly explored this interaction in more detail.

## Data Availability

All data produced in the present study are available upon reasonable request to the authors.

## Supplementary

**Figure S1:**
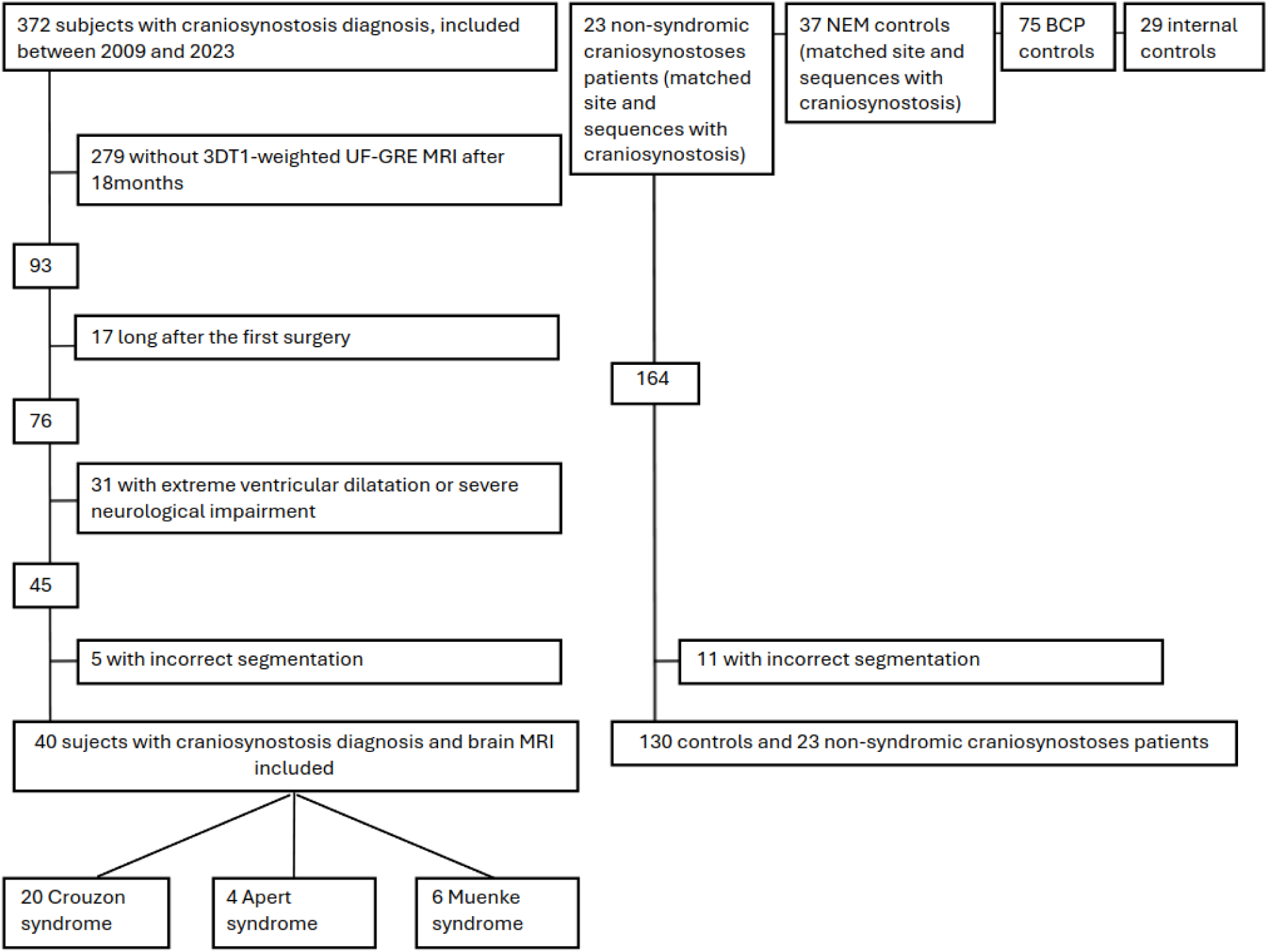
Flowchart showing the composition of craniosynostosis and control groups included in the analyses.

**Table S1.** Tests evaluating the mean deviations from control for site-effect validation. Mean residual differences (cm³) between subgroups of controls from the NEM site and the normative model in each subtest, obtained through 4-fold cross-validation. Values are shown as mean residual difference [95% confidence interval], with p adjusted using the FDR-BY method.

| Volume (cm<sup>3</sup>) | Fold 1 | Fold 2 | Fold 3 | Fold 4 |
|---|---|---|---|---|
| Central | 1.29<br>[-4.43,9.86]<br>(1) | -0.67<br>[-6.14,7.46]<br>(1) | -2.71<br>[-8.51,5.89]<br>(1) | -2.98<br>[-7.84,6.13]<br>(1) |
| Frontal | 3.01<br>[-10.07,29.14]<br>(1) | 9.11<br>[-3.23,3.39]<br>(1) | -16.38<br>[-27.90,10.37]<br>(1) | -2.52<br>[-14.14,24.03]<br>(1) |
| Parietal | 4.61<br>[-8.41,14.17]<br>(1) | 2.92<br>[-10.45,11.70]<br>(1) | 7.47<br>[-5.48,16.36]<br>(1) | 5.41<br>[-6.98,14.85]<br>(1) |
| Temporal | 0.05<br>[-11.74,15.47]<br>(1) | -6.76<br>[-18.48,7.99]<br>(1) | -4.07<br>[-15.64,10.98]<br>(1) | -5.55<br>[-17.08,9.35]<br>(1) |
| Occipital | -10.81<br>[-9.18,9.79]<br>(1) | -10.56<br>[-8.54,9.25]<br>(1) | -13.54<br>[-11.29,6.70]<br>(1) | -9.21<br>[-6.67,11.56]<br>(1) |
| Brainstem | -0.45<br>[-0.74,1.62]<br>(1) | 0.20<br>[-0.03,2.26]<br>(1) | -1.39<br>[-1.63,0.79]<br>(1) | -0.57<br>[-0.82,1.52]<br>(1) |
| Lateral ventricles | 0.76<br>[-8.67,3.25]<br>(1) | -1.55<br>[-10.96,0.89]<br>(1) | 2.05<br>[-7.45,4.45]<br>(1) | -2.11<br>[-11.44,0.51]<br>(1) |
| Cerebellum | -6.16<br>[-11.80,3.52]<br>(1) | -1.68<br>[-7.65,7.33]<br>(1) | -3.00<br>[-9.16,6.11]<br>(1) | 0.15<br>[-6.06,9.14]<br>(1) |
| TBV | -0.85<br>[-36.29,74.35]<br>(1) | -4.93<br>[-39.05,63.89]<br>(1) | -29.80<br>[-63.04,44.47]<br>(1) | -15.23<br>[-46.09,59.79]<br>(1) |

**Table S2.** Results of statistical tests assessing normality (Shapiro–Wilk) and homoscedasticity (Breusch–Pagan, Levene) of residuals from the normative age-effect models in the control population: mean residuals difference (p adjusted using the FDR-BY method)

| Volume (p-values) | Shapiro-Wilk W statistic | Breusch-Pagan LM statistic | Levene F statistic (5 age groups) |
|---|---|---|---|
| Central | 0.99 (1) | 3.09 (0.90) | 0.96 (1) |
| Frontal | 0.98 (1) | 1.53 (1) | 0.59 (1) |
| Parietal | 0.99 (1) | 0.24 (1) | 0.65 (1) |
| Temporal | 0.99 (1) | 3.09 (1) | 1.93 (1) |
| Occipital | 0.99 (1) | 3.56 (1) | 1.16 (1) |
| Brainstem | 0.99 (1) | 1.49 (1) | 0.37 (1) |
| Lateral ventricles | 0.83 (10<sup>-9</sup>) | 0.71 (1) | 0.22 (1) |
| Cerebellum | 0.99 (1) | 3.08 (1) | 1.35 (1) |
| TBV | 0.99 (0.80) | 3.40 (0.58) | 0.83 (1) |

**Figure S2:**
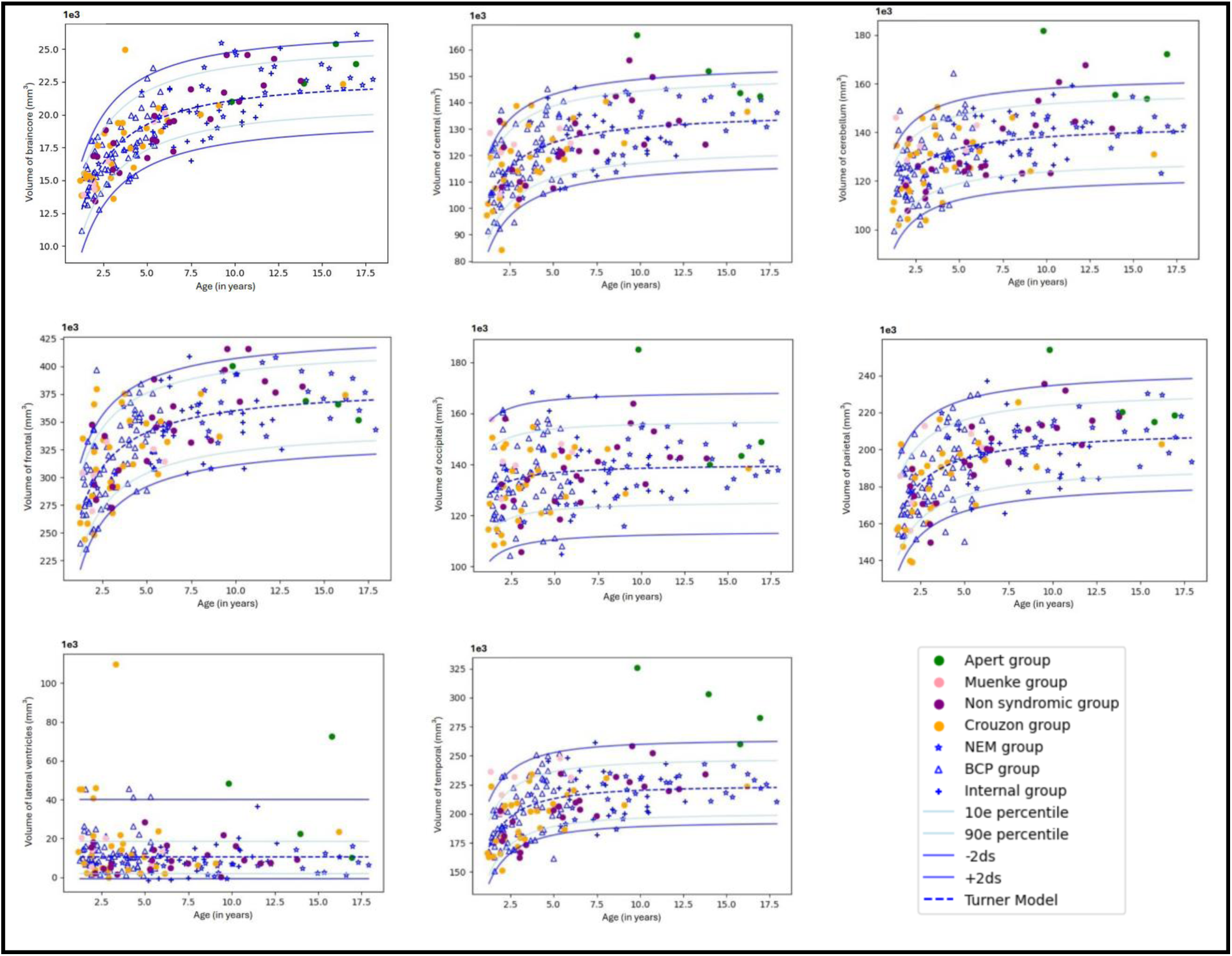
Age effect charts for regional brain volumes in the patient groups (colored dots: green for Apert syndrome, pink for Muenke syndrome, purple for non-syndromic, orange for Crouzon syndrome) compared with normative curves from the control group. Controls are represented with different symbols according to their origin: stars for the NEM group (*), triangles for the BCP group (Δ), and crosses for the internal group (+). The fitted model curve for controls is shown as a blue dotted line. The other blue lines represent the 10^th^ and 90^th^ percentiles, as well as ±2 standard deviations of the normative distribution.

**Table S3.** Tests evaluating the distribution deviations from control of the age effect. p-values (FDR-BY adjusted) from statistical tests comparing the distribution of regional brain volumes between each patient group and the control group in the age-effect analysis.

| Volume (p-values) | Crouzon group<br>n=30 | Apert group<br>n=4 | Muenke group<br>n=6 | Non-syndromic group<br>n=23 |
|---|---|---|---|---|
| Central | 1 | 0.21 | 1 | 1 |
| Frontal | 1 | 1 | 1 | 1 |
| Parietal | 1 | 0.818 | 1 | 1 |
| Temporal | 1 | **0.0014** | **0.052** | 1 |
| Occipital | 1 | 0.818 | 1 | 1 |
| Brainstem | 1 | 1 | 1 | 1 |
| Lateral ventricles | 0.21 | **0.042** | 1 | 1 |
| Cerebelum | 1 | **0.0016** | 1 | 1 |
| TBV | 1 | **0.0012** | 1 | 1 |

**Figure S3:**
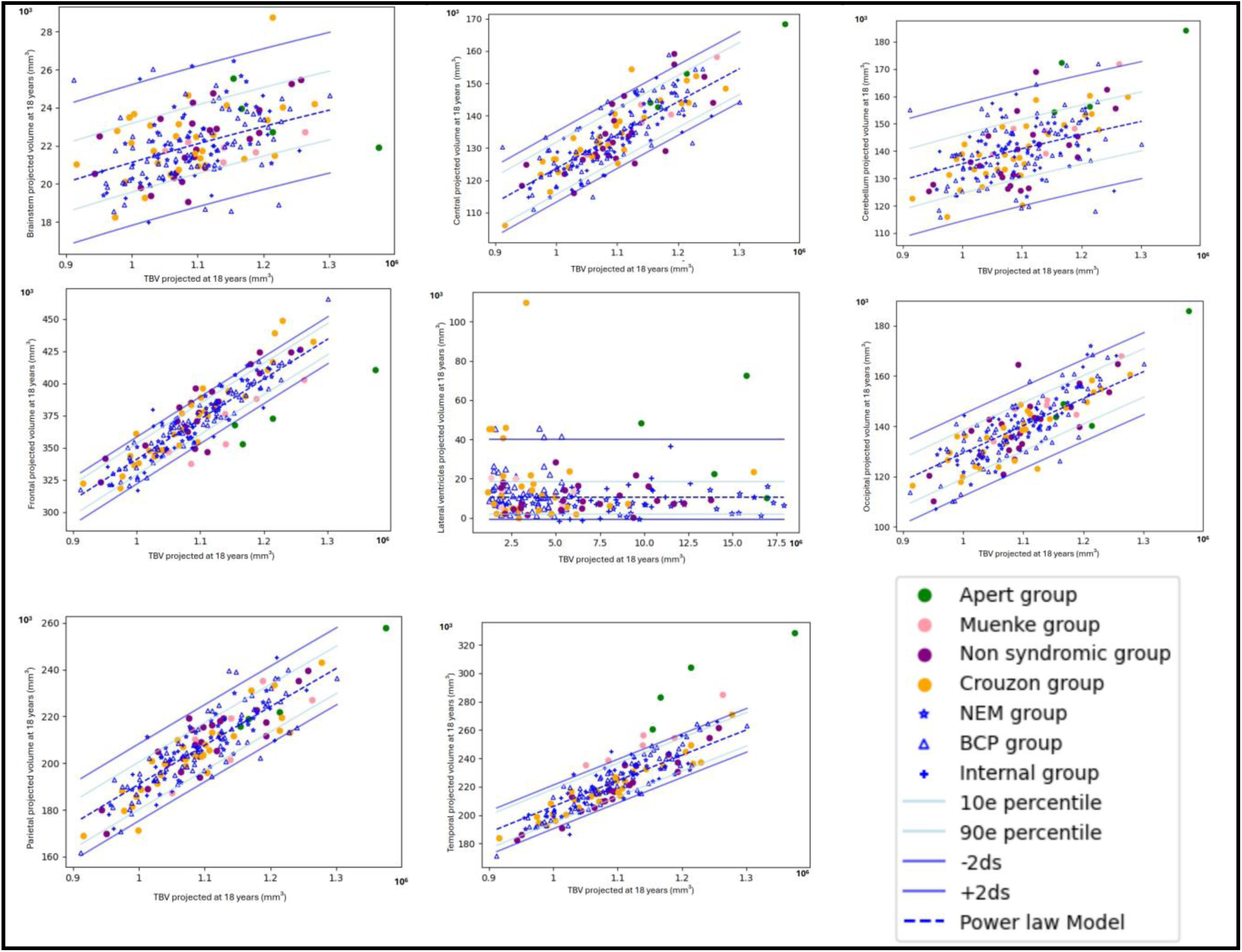
Graphs of normative curves from the power law model adjusted for controls (in blue), with patient groups shown as colored dots (green for Apert syndrome, pink for Muenke syndrome, purple for non-syndromic, orange for Crouzon syndrome). Controls are represented with different symbols according to their origin: stars for the NEM group (*), triangles for the BCP group (Δ), and crosses for the internal group (+). The fitted model curve for controls is shown as a blue dotted line. The other blue lines represent the 10^th^ and 90^th^ percentiles, as well as ±2 standard deviations of the normative distribution.

**Table S4.** Tests evaluating the distribution deviations from control in static scaling. p-values (FDR-BY adjusted) from statistical tests comparing the distribution of regional brain volumes projected at 18 years between each patient group and the control group after adjusting for total brain volume (TBV) projected at 18 years.

| Volume (p-values) | Crouzon group<br>n=30 | Apert group<br>n=4 | Muenke group<br>n=6 | Non-syndromic group<br>n=23 |
|---|---|---|---|---|
| Central | 1 | 0.93 | 1 | 0.73 |
| Frontal | 1 | **0.0013** | **0.021** | 0.33 |
| Parietal | 1 | 1 | 1 | 0.96 |
| Temporal | 1 | **0.0013** | **10<sup>-5</sup>** | 0.73 |
| Occipital | 1 | 0.56 | 1 | 1 |
| Brainstem | 1 | 0.93 | 1 | 1 |
| Lateral ventricles | 0.48 | 0.56 | 1 | 1 |
| Cerebellum | 1 | **0.044** | 1 | 0.54 |

**Figure S4:**
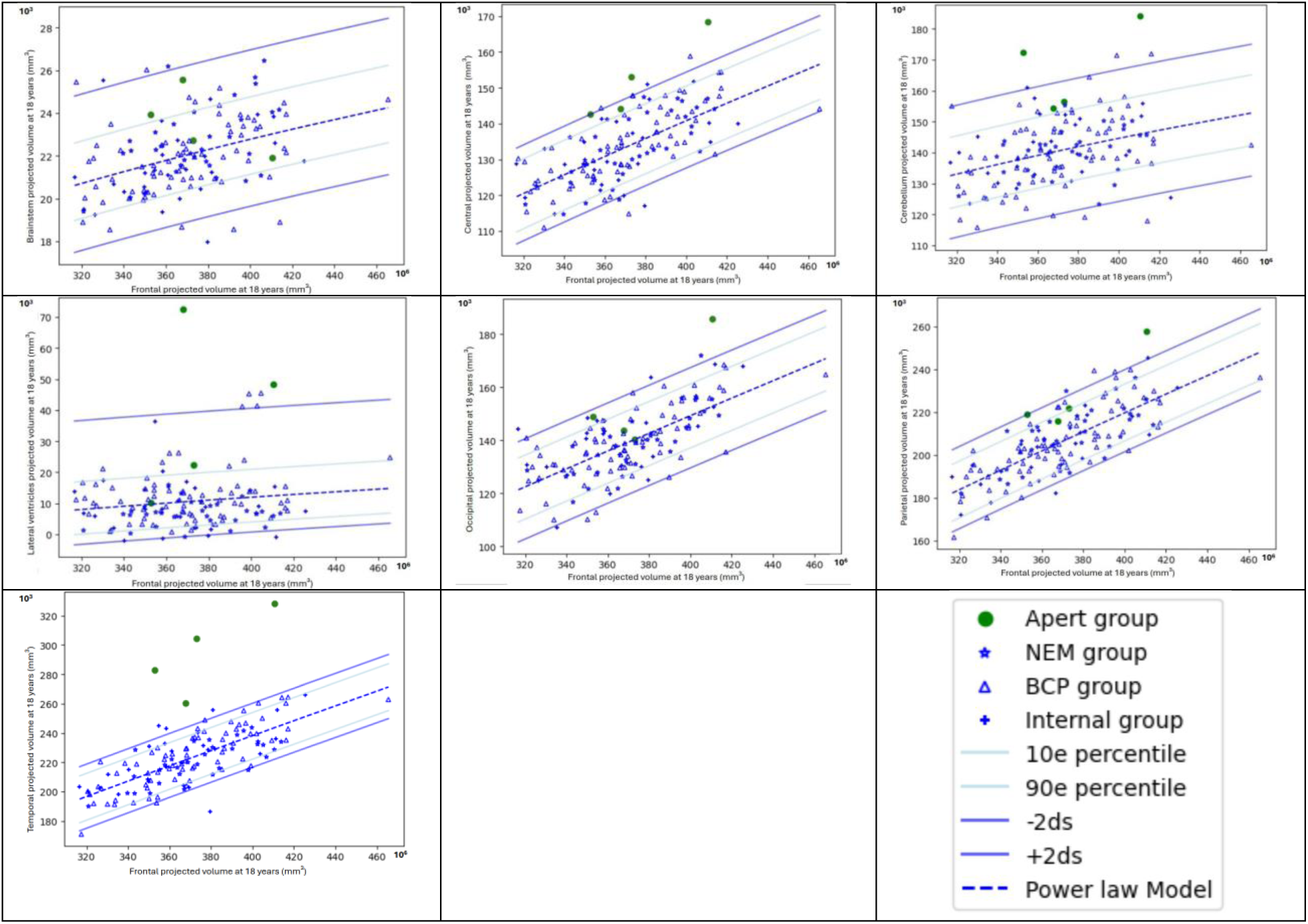
Graphs of normative curves from the power law model adjusted for controls (in blue) in relation to frontal volume, with patient groups shown as colored dots (green for Apert syndrome). Controls are represented with different symbols according to their origin: stars for the NEM group (*), triangles for the BCP group (Δ), and crosses for the internal group (+). The fitted model curve for controls is shown as a blue dotted line. The other blue lines represent the 10^th^ and 90^th^ percentiles, as well as ±2 standard deviations of the normative distribution.

**Figure S5.**
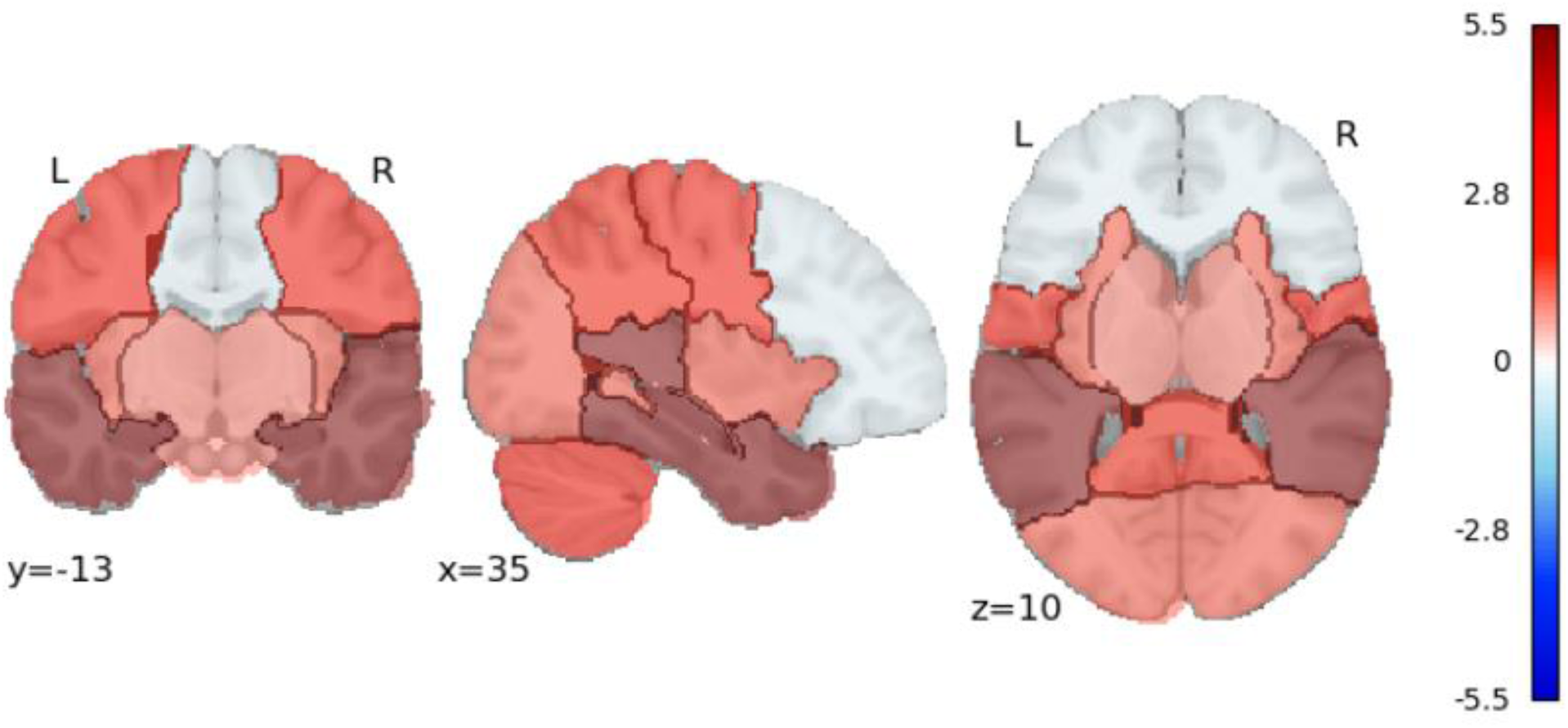
Normative pattern and extent of regional oversizing in Apert syndrome when frontal volume, that is normal for age, is taken as a reference.

**Table S5.** Comparison of regional brain volumes (cm³) projected at age 18years between preoperative and postoperative patients with Crouzon syndrome. Values are presented as mean ± standard deviation. p-values are from Welch’s t-test.

| Volume (cm<sup>3</sup>) | Mean ± SD<br>preoperative | Mean ± SD<br>postoperative | t(df) | p |
|---|---|---|---|---|
| **Central** | 131.39±11.31 | 137.27±9.40 | -1.38(11.8) | 0.19 |
| **Frontal** | 365.52±33.29 | 385.22±35.33 |  | 0.22 |
| **Parietal** | 198.97±16.82 | 211.88±14.76 | -1.96(11.2) | 0.075 |
| **Temporal** | 216.17±20.07 | 227.04±11.94 |  | 0.095 |
| **Occipital** | 134.75±12.43 | 142.95±10.20 | -1.77(12.0) | 0.10 |
| **Brainstem** | 22.57±2.19 | 22.76±1.20 |  | 0.77 |
| **Lateral ventricles** | 18.34±24.13 | 14.99±15.31 | 0.44(16.0) | 0.67 |
| **Cerebellum** | 137.48±11.58 | 143.32±11.37 |  | 0.26 |
| **TBV** | 1066.48±87.84 | 1124.15±73.35 | -1.74(11.8) | 0.11 |

## Data and Code Availability

The data and code supporting the findings of this study are available from the corresponding authors upon reasonable request.

## Funding

This work was supported by the “Agence Nationale de la Recherche,” “Investissements d’Avenir” program (ANR-10-IAHU-01) by France 2030 grant “Face4Kids” (ANR-21-PMRB-0004), and IRDCE (a grant from the French government managed by the Agence Nationale de la Recherche as part of the France 2030 program, ANR-23-IAHU-0010).

## Declaration of Competing Interests

The authors declare that they have no known competing financial interests or personal relationships that could have appeared to influence the work reported in this paper.

## Notes

### Competing Interest Statement

The authors have declared no competing interest.

### Author Declarations

Ethics committee of Necker-Enfants Malades University Hospital gave ethical approval for this work.

